# Early Stage Prediction of US County Vulnerability to the COVID-19 Pandemic

**DOI:** 10.1101/2020.04.06.20055285

**Authors:** Mihir Mehta, Juxihong Julaiti, Paul Griffin, Soundar Kumara

## Abstract

**Importance:** The rapid spread of COVID-19 means that government and health services providers have little time to plan and design effective response policies. It is therefore important to rapidly provide accurate predictions of how vulnerable geographic regions such as counties are to the spread.

**Objective:** Developing county level prediction around near future disease movement for COVID-19 occurrences using publicly available data.

**Design:** Original Investigation; Decision Analytical Model Study for County Level COVID-19 occurrences using data from March 14-31, 2020.

**Setting:** Disease spread prediction for US counties.

**Participants:** All US county level granularity based on data fused from multiple publicly available sources inclusive of health statistics, demographics, and geographical features.

**Exposure(s) (for observational studies):** Daily county level reported COVID-19 occurrences from March 14-31, 2020.

**Main Outcome(s) and Measure(s):** We developed a 3-stage model to quantify, firstly the probability of COVID-19 occurrence for unaffected counties using XGBoost classifier and secondly, the number of potential occurrences of a county via XGBoost regression. Thirdly, these results are combined to compute the county level risk. This risk is then used as an estimated after-five-day-vulnerability of the county.

**Results:** Using data from March 14-31, 2020, the model shows a sensitivity over 71.5% and specificity over 94%.

**Conclusions and Relevance:** We found that population, population density, percentage of people aged 70 or greater and prevalence of comorbidities play an important role in predicting COVID-19 occurrences. We found a positive association between affected and urban counties as well as less vulnerable and rural counties. The developed model can be used for identification of vulnerable counties and potential data discrepancies. Limited testing facilities and delayed results introduces significant variation in reported cases and produces a bias in the model.

**Trial Registration:** Not Applicable

**Key Points:** *Question:* What are key factors that define the vulnerability of counties in the US to cases of the COVID-19 virus?

*Findings:* In this epidemiological study based on publicly available data, we develop a model that predicts vulnerability to COVID-19 for each US county in terms of likelihood of going from no documented cases to at least one case within five days and in terms of number of occurrences of the virus.

*Meaning:* Predicting county vulnerability to COVID-19 can assist health organizations to better plan for resource and workforce needs.

## Introduction

The continued spread of confirmed cases of COVID-19, absence of a vaccine, limited resources for testing and assisting people with confirmed cases have presented a great challenge for our public health and healthcare provider systems. To this point, nonpharmaceutical interventions such as social distancing are the only effective mitigation measures. The rapid spread of the disease means that government and health services have very little time to plan and design effective response policies such as resource and workforce planning. Accurately predicting the near future COVID-19 spread at sufficient granularity would provide these organization with better information and time to appropriately plan and respond.

We have developed a three-stage machine learning model to estimate COVID-19 spread outcomes at the US county level. In the first stage, we estimate the probability that a county has at least one confirmed COVID-19 case. In the second stage, we estimate the number of COVID-19 occurrences given that county has at least one case. Finally, we combine the results from the two stages to estimate those counties that have the greatest and least vulnerability for changes in disease prevalence for the next five-day period.

There has been significant epidemiological work for previous coronavirus pandemics such as MERS and SARS.^1^ For example, Badawi et al.^2^ performed systematic analysis of prevalence of comorbidities in MERS using data from 12 studies and found that diabetes and hypertension were present in 50% of the cases. Matsuyama et al.^3^ systematically reviewed studies involving laboratory confirmed MERS cases to measure both the risk of admission to the Intensive Care Unit (ICU) and death. They compared risks by age, gender and underlying comorbidities. Park et al.^4^ reviewed characteristics and associated risks factors of MERS. Bauch et al.^5^ surveyed SARS modeling literature focused on understanding the basic epidemiology of the disease and evaluating control strategies. Surveyed SARS models varied in the terms of population studied and geographical characteristics.^6,7^ Different designs were used for SARS modeling consisting of deterministic compartmental models^7^, stochastic compartmental models^6^, a combination of stochastic and deterministic compartmental models^8^, discrete-time models^9^, logistics curve fitting models^10^, contact network models^11^ and likelihood-based models.^12^ Studies associated with risk factors for SARS^13^ and MERS^3,14–20^ have found an association between comorbidities and infected cases.

MERS and SARS epidemiological modeling has been done at different granularities such as the country^21,22^, specific region^23^, and case clusters.^6^ Given the much broader reach of COVID-19 compared to MERS and SARS, it is very important to predict at a sufficiently high level of granularity. This is particularly important since previous studies have shown that there is considerable heterogeneity in space, transmissibility and susceptibility.^5^ Our approach is developed at county level with inclusion of a variety of health statistics, demographics and geographical features of counties. Further, we use publicly available data so that any organization could use the model. To the best of our knowledge, no work has been done to predict near future infection risk at the county level using the combination of health statistics, demographics and geographical features of counties.

## Methods

### Study Design and Population

We performed an epidemiological study at the US county level using publicly available data to develop a machine learning predictive model. Data analysis was performed from February 15, 2020, to April 3, 2020. The study was reviewed by the Penn State Integrated Research Ethics Board and deemed exempt because it was a deidentified, secondary data analysis. This study followed the Strengthening the Reporting of Observational Studies in Epidemiology (STROBE) reporting guideline.^24^

### Data Sources

We used US Census data to obtain county level population statistics for age, gender and density. ^25,26^ We obtained county level data for diagnosed adult diabetics percentage and cancer crude rate statistics from the Center for Disease Control and Prevention (CDC).^27,28^ We used county level hypertension estimates and chronic respiratory disease mortality rates from the Global Heath Data Exchange (GHDx)^29,30^, provided by the Institute for Health Metrics and Evaluation. We obtained the centroids for each county from ArcGIS.^31^ Finally, we obtained US Census Cartographic Boundary files for each county in JSON format^32^ and county level COVID-19 daily occurrences data (confirmed cases) from NYTimes GitHub page.^33,34^

### Outcomes

There are three primary outcomes for our predictive model: i) the probability that a county has at least one confirmed case of COVID-19, which we define as a positive instance, ii) the number of confirmed COVID-19 cases within a county, which we define as occurrences, and iii) vulnerability of the county.

### Covariates

Previous studies have shown angiotensin-converting enzyme 2 (ACE2) facilitates the infection of COVID-19^35–37^, and that patients with diabetes, hypertension and cardiovascular diseases have an increased expression of ACE2.^35^ County population factors such as density, age, and sex have a significant impact on the spread of an epidemic.^38^ Cancer and chronic respiratory diseases have also been shown to increase mortality risk for COVID-19.^39^

The dataset used for our three-stage model contains correlated variables. For example, diabetes and hypertension prevalence, cancer crude rate and old population. Additionally, the underlying relationship between variables was assumed to be non-linear. For such cases the literature supports ^40–47^ using gradient tree boosting and deep learning methods for better prediction results.

### Statistical Analysis

#### Developing the Prediction Model

In order to predict COVID-19 outcomes, we divided the problem into three stages. In the first stage, we formulated a binary classification problem that included both positive and negative instances. We developed an XGBoost^48^ classifier model to learn from the data. We divided the dataset into training and testing in 80-20 proportions for each class. We tuned the hyperparameters of the model using the Hyopt package.

In the second stage, we formulated an XGBoost regression model that included data only for positive instances with number of occurrences as the response. As in the case for the first stage, we divided data into training and testing sets in 80-20 proportions and used the Hyopt package for hyperparameter tuning.

In the last stage, we combined results from the first two stages and calculated the expected occurrences for counties as a measure of county vulnerability. For the calculation of expected occurrences, we multiplied the probability of county belonging to the positive instances derived using the classification model, with potential occurrences the same county will have if it becomes a positive instance derived using the regression model.

#### Evaluating the Prediction Model

Area under the receiver operating characteristic curve (AUC) and accuracy are used as the criteria to evaluate the classification model (the first stage of the model). The root mean squared error (RMSE) is used as the criteria to evaluate the regression model (the second stage of the model). The final stage of the model-vulnerability was assessed by examining the sensitivity and specificity of the prediction.

## Results

The variable importance for the overlapping predictors between the final classification and regression models for March 16^th^ is shown in Figure 1. Total population (TOT_POP) was the most important variable for both the classification and regression models. Other important variables included population density, longitude, hypertension prevalence, chronic respiratory mortality rate, cancer crude rate, and diabetes prevalence. Latitude (we use this to identify neighboring counties and the presence or absence of positive class in the neighborhood) and percentage of populations older than 70 years were found to be the least important features of those considered, though still played a role.

**Figure 1:**
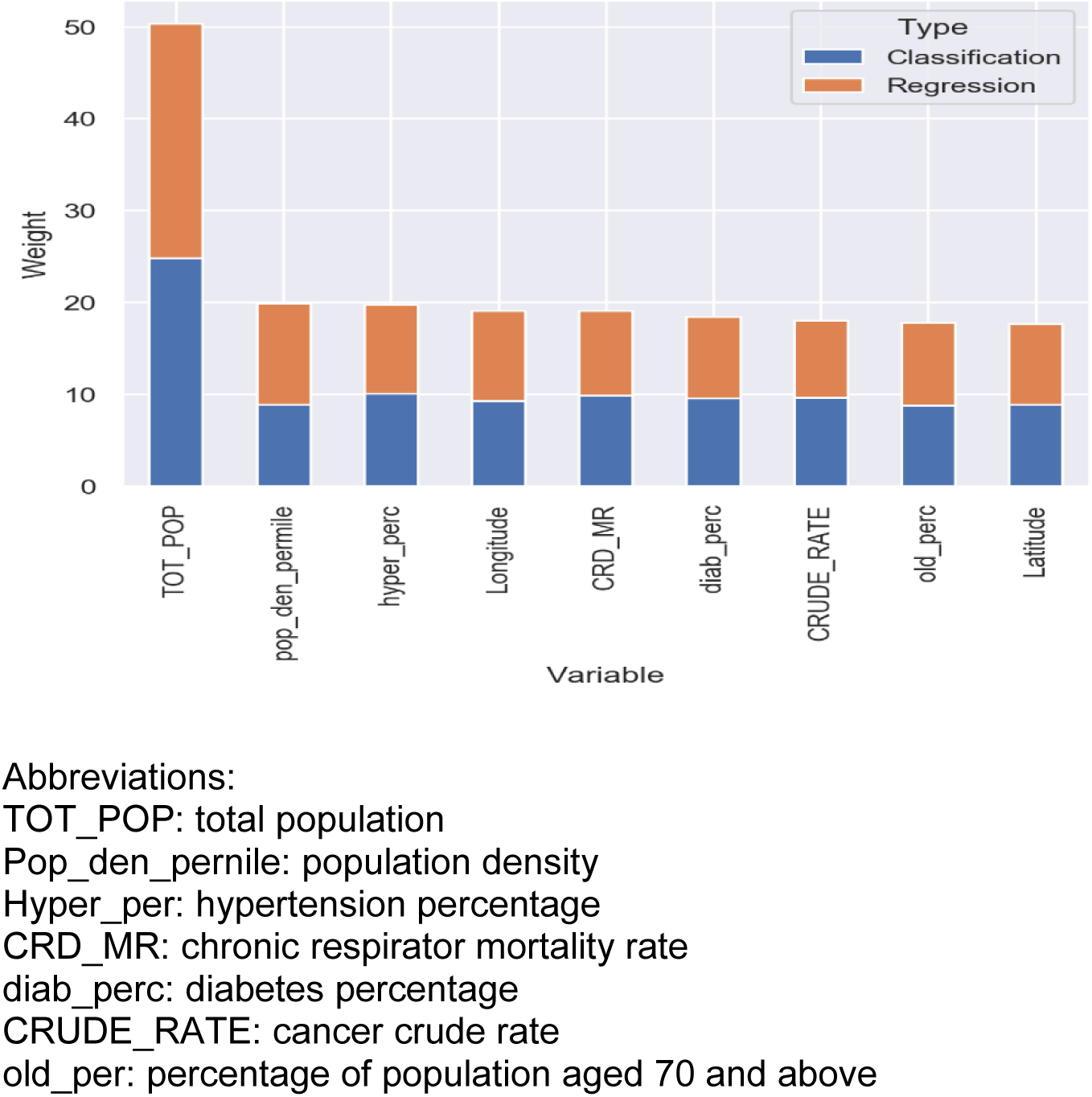
Variable Importance for the Classification and Regression Models.

Figure 2 shows a map of the USA with the predicted probability of being a positive instance for each county in the USA as a color gradient. County level statistics can be viewed by moving the cursor of the county of interest. The example of New York County as of March 14^th^ is shown in the Figure 2.

**Figure 2:**
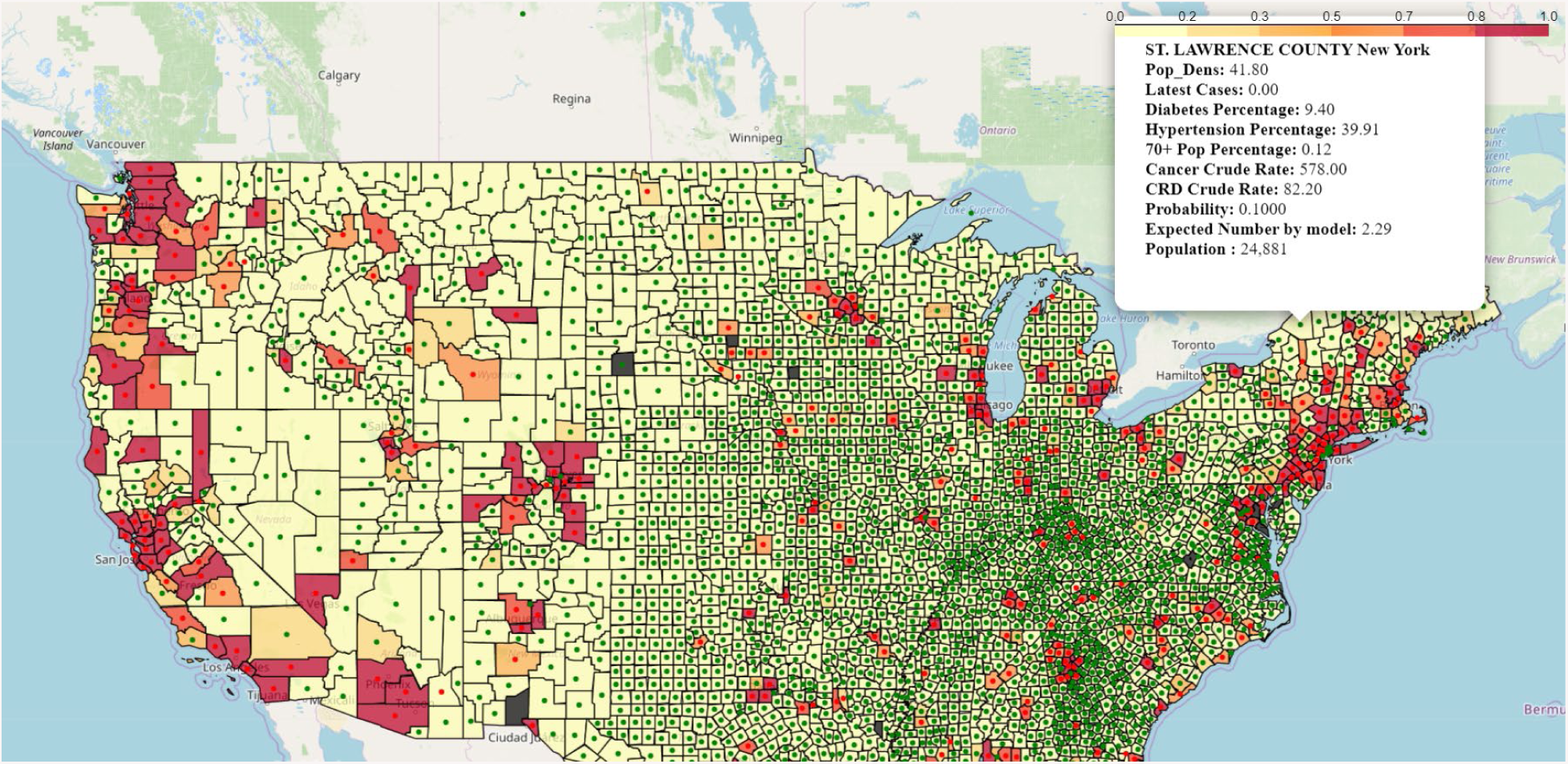
County Level COVID-19 Vulnerability Map for the US.

Accuracy and AUC for the first stage model is shown in Table 1. Predictions of the model for all US counties are consistent over the 18 days with little variation in AUC and accuracy values. Similarly, RMSE for the second stage model for all US counties is presented in Table e1.

**Table 1:** XGBoost Classification Training and Testing Details.

| Dataset | Metrics | Mean | Min | Max | Standard Deviation | Number of Days |
| --- | --- | --- | --- | --- | --- | --- |
| Test | Accuracy | 83% | 77% | 92% | 5% | 18 |
|  | AUC | 78% | 71% | 83% | 3% | 18 |
| Train | Accuracy | 94% | 82% | 100% | 5% | 18 |
|  | AUC | 91% | 80% | 100% | 6% | 18 |

The sensitivities and specificities for the vulnerability predictions for the three-stage model trained on data from March 14^th^ to March 26^th^ are shown in Tables 2 and 3. The values are given for each day. The sensitivity (Table 2) is given by percentage of counties that had no confirmed cases but were identified as being among the 5% most vulnerable had at least one confirmed COVID-19 case five days later. The specificity (Table 3) is given by the percentage of counties identified as being among the 10% least vulnerable with no confirmed cases that still had no confirmed cases five days later.

**Table 2:**
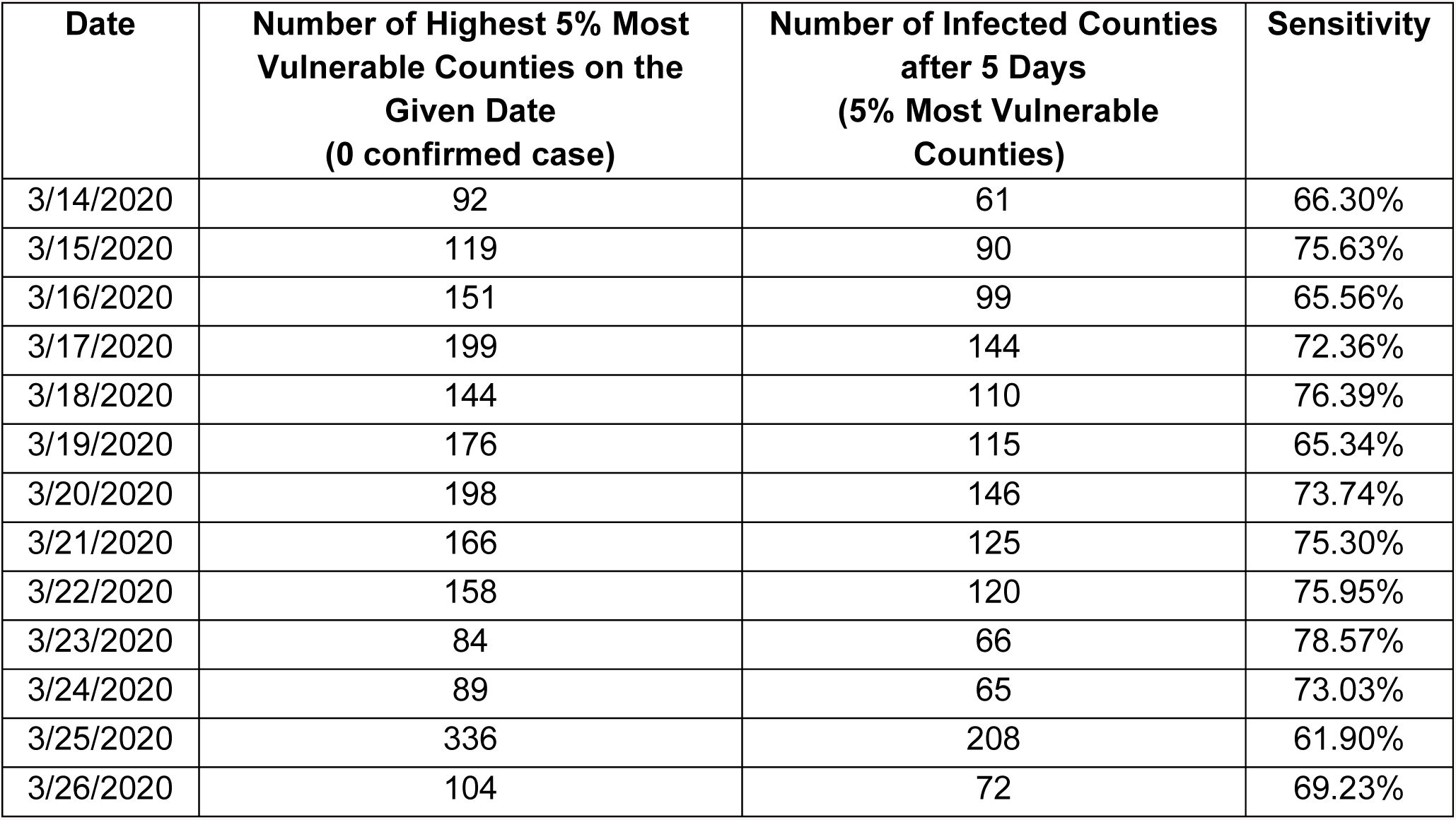
Sensitivity of the three-stage Model.

| **Date** | **Number of Highest 5% Most Vulnerable Counties on the Given Date**<br>**(0 confirmed case)** | **Number of Infected Counties after 5 Days**<br>**(5% Most Vulnerable Counties)** | **Sensitivity** |
| --- | --- | --- | --- |
| 3/14/2020 | 92 | 61 | 66.30% |
| 3/15/2020 | 119 | 90 | 75.63% |
| 3/16/2020 | 151 | 99 | 65.56% |
| 3/17/2020 | 199 | 144 | 72.36% |
| 3/18/2020 | 144 | 110 | 76.39% |
| 3/19/2020 | 176 | 115 | 65.34% |
| 3/20/2020 | 198 | 146 | 73.74% |
| 3/21/2020 | 166 | 125 | 75.30% |
| 3/22/2020 | 158 | 120 | 75.95% |
| 3/23/2020 | 84 | 66 | 78.57% |
| 3/24/2020 | 89 | 65 | 73.03% |
| 3/25/2020 | 336 | 208 | 61.90% |
| 3/26/2020 | 104 | 72 | 69.23% |

**Table 3:** Specificity of the three-stage Model.

| **Date** | **Number of Top 10% Least Vulnerable Counties on the Date**<br>**(0 confirmed case)** | **Number of Counties with 0 case after 5 Days**<br>**(Top 10% Least Vulnerable Counties)** | **Specificity** |
| --- | --- | --- | --- |
| 3/14/2020 | 276 | 274 | 99.28% |
| 3/15/2020 | 282 | 276 | 97.87% |
| 3/16/2020 | 46 | 44 | 95.65% |
| 3/17/2020 | 313 | 304 | 97.12% |
| 3/18/2020 | 297 | 281 | 94.61% |
| 3/19/2020 | 214 | 198 | 92.52% |
| 3/20/2020 | 295 | 266 | 90.17% |
| 3/21/2020 | 312 | 291 | 93.27% |
| 3/22/2020 | 15 | 14 | 93.33% |
| 3/23/2020 | 310 | 289 | 93.23% |
| 3/24/2020 | 303 | 270 | 89.11% |
| 3/25/2020 | 214 | 197 | 92.06% |
| 3/26/2020 | 231 | 218 | 94.37% |

The dataset is comprised of 37% urban and 63% rural counties based on the urban and rural county definition for year 2013.^49^ In order to determine if there is an association between urbanicity and vulnerability, we performed a set of one-sided t-tests. The null hypothesis - the 10% least vulnerable counties would have the same proportion of rural counties as the actual proportion of rural counties in the dataset - was rejected for every day from March 14^th^ to March 26^th^. Additionally, the null hypothesis - the actual positive instances counties would the same proportion of urban counties as the actual proportion of urban counties in the dataset - was also rejected for every day over the analysis period. It can therefore be concluded that there is a positive association between urban and most vulnerable counties as well as rural and least vulnerable counties. The continuous decreasing trend in the confidence interval of the urban counties proportion estimate within actual positive instance counties can be used to infer that COVID-19 is propagating from urban counties to rural counties.

## Discussion

We developed a three-stage machine learning model using publicly available data to predict the five-day vulnerability of a US county. The model estimates the likelihood and impact that a county with no documented COVID-19 cases will have within a five-day period and using them, vulnerability prediction for a county is made. Using data from March 14^th^ to Marth 31^st^, 2020, the model showed a sensitivity over 71.5% and specificity over 94%. We found a positive association between affected counties and urban counties as well as top 10% least vulnerable counties and rural counties. Further, counties with higher population density, a greater percentage of 70 years of above age people, higher diabetes, cardiac illness and respiratory diseases prevalence are more vulnerable to COVID-19 than their counterparts.

Our model serves multiple purposes. First, it can help in identifying potentially vulnerable counties. This prediction would be a vital component in managing COVID-19 spread by providing vulnerability information based on the likelihood and magnitude of change within five days. That can help health organizations to plan effectively for management of hospital resources and workforce, rapid response teams, and COVID testing kits and testing locations. In addition, there are multiple counties with limited testing facilities, and with current swab-based testing, it takes multiple days to get the results. Thus, occurrences associated with each county fluctuate rapidly daily.

There are multiple limitations to our work. First, there are several predictors that we did not include in the model that have known associations with COVID-19. However, one of our goals was to make sure that any organization could use our model by only including data that is publicly available. Second, our analysis (Table e2) found that there is an increasing trend for the coefficient of variation (CV) for occurrences associated with positive instances counties.

Note that CV is a proxy for economic inequality.^50–53^ Hence, there is a bias in the response variable, which can reduce the accuracy of the prediction. As testing facilities improve in terms of numbers and efficiency, this bias would be minimized and would be reflected in the model. Given this point, it would useful to look at top riskiest and top safest counties predicted by MJK model and examine for potential data discrepancies. Finally, additional feature engineering and stacking methods can be utilized to enhance the prediction capabilities of existing models.

Our work uses open source programming and publicly available data. We will make the full dataset, sample modeling and result outputs available with instructions for use soon on: https://github.com/mihirpsu/covid_19

## Data Availability

Our work uses open source programming and publicly available data. We will make the full dataset, sample modeling and result outputs available with instructions for use soon on:
https://github.com/mihirpsu/covid_19

## Supplementary Materials

**Table e1:** XGBoost Regression Training and Testing Details.

| Dataset | Metrics | Mean | Min | Max | Standard Deviation | Number of Days |
| --- | --- | --- | --- | --- | --- | --- |
| Test | RMSE | 117.10 | 7.13 | 372.97 | 105.00 | 18 |
| Train | RMSE | 14.02 | 1.19 | 48.59 | 14.28 | 18 |

**Table e2:** COVID-19 Daily Positive Occurences Descriptive Statistics.

| Date | Mean | Standard Deviation | Coefficient of Variation |
| --- | --- | --- | --- |
| 3/14/2020 | 7.01 | 25.25 | 3.60 |
| 3/15/2020 | 7.63 | 26.44 | 3.46 |
| 3/16/2020 | 8.53 | 29.39 | 3.44 |
| 3/17/2020 | 9.66 | 33.76 | 3.49 |
| 3/18/2020 | 10.96 | 38.91 | 3.55 |
| 3/19/2020 | 12.98 | 49.54 | 3.81 |
| 3/20/2020 | 14.85 | 61.98 | 4.17 |
| 3/21/2020 | 17.56 | 78.73 | 4.48 |
| 3/22/2020 | 20.93 | 103.22 | 4.93 |
| 3/23/2020 | 24.96 | 135.00 | 5.41 |
| 3/24/2020 | 28.21 | 162.05 | 5.74 |
| 3/25/2020 | 30.82 | 181.81 | 5.90 |
| 3/26/2020 | 36.29 | 218.46 | 6.02 |
| 3/27/2020 | 41.86 | 255.72 | 6.11 |
| 3/28/2020 | 47.43 | 288.14 | 6.07 |
| 3/29/2020 | 52.69 | 322.05 | 6.11 |
| 3/30/2020 | 58.01 | 354.68 | 6.11 |
| 3/31/2020 | 64.93 | 395.28 | 6.09 |

## Supplementary Results

Table e3 shows a sample list of negative instance counties as of March 14^th^. The 3-stage model predicted them in the top 5% riskiest counties additional to infected counties. All these sample counties were identified as positive instances on March 19^th^. Similarly, Table e4 shows a sample list of negative instance counties as of March 14^th^. The 3-stage model predicted them as top 10% safest counties. All these sample counties continued to be negative instances on March 19^th^ as shown in the table.

**Table e3:** Samples of Counties from the Top 5% Riskiest Counties with Negative Instances.

| State | County | Number of cases on March 14th | Number of cases on March 19<sup>th</sup> |
| --- | --- | --- | --- |
| Florida | Leon | 0 | 3 |
| Illinois | Will | 0 | 11 |
| Maine | York | 0 | 3 |
| Massachusetts | Plymouth | 0 | 5 |
| Minnesota | Washington | 0 | 3 |
| New York | Erie | 0 | 27 |
| Texas | Denton | 0 | 9 |
| Wisconsin | Kenosha | 0 | 4 |

**Table e4:** Samples of Counties from the Top 10% Safest Counties.

| State | County | Number of cases on March 14th | Number of cases on March 19th |
| --- | --- | --- | --- |
| Georgia | Glascok | 0 | 0 |
| Kansas | Smith | 0 | 0 |
| Kentucky | Hickman | 0 | 0 |
| Mississippi | Issaquena | 0 | 0 |
| New Mexico | Catron | 0 | 0 |
| North Dakota | Emmons | 0 | 0 |
| Texas | Jack | 0 | 0 |
| Texas | Sutton | 0 | 0 |

